# HealthFormer: Dual-level time-aware Transformers for irregular electronic health record events

**DOI:** 10.64898/2026.03.25.26349262

**Authors:** Péter Kőrösi-Szabó, Gábor Kovács, Botond Forrai, Judit Laki, Tamás Kováts, Miklós Szócska, Adrián Csiszárik

**Affiliations:** HUN-REN Alfréd Rényi Institute of Mathematics, Budapest, Hungary; Data-Driven Health Division of National Laboratory for Health Security, Health Services Management Training Centre, Semmelweis University, Budapest, Hungary; Mossavar-Rahmani Center for Business and Government, Harvard University, Kennedy School of Government

**Keywords:** electronic health records, self-supervised pretraining, hierarchical Transformer, time-aware attention, representation learning

## Abstract

Longitudinal electronic health records (EHRs) form irregular event sequences that mix multiple clinical coding systems and care settings. Learning transferable patient representations requires modeling both within-encounter code composition and long-range temporal dependencies. We aim to develop a pretraining framework that preserves event structure and explicitly uses elapsed time, while remaining straightforward to fine-tune for new supervised endpoints without task-specific feature engineering. We propose HealthFormer, a dual-level Transformer for event-centric EHR modeling. An *Intra-Event Encoder* aggregates heterogeneous domain tokens within each typed clinical event into an event embedding via code-specific embedding modules and attention pooling. Event embeddings are combined with a *Date Encoder* and a continuous-time attention bias based on attention with linear biases (ALiBI) inside an *Inter-Event Encoder*. We pretrain on Hungarian national administrative health records from a large-scale nationwide longitudinal cohort (spanning millions of individuals over a decade) using multi-task self-supervision with (i) per-domain masked token prediction (masked language modeling, MLM), (ii) event-type prediction under full-event masking (Event-level MLM), (iii) next-event type prediction, and (iv) time-to-next-event (Δ*t*) regression. Pretraining induces hierarchy-consistent organization in learned diagnosis (ICD-10) embedding geometry conducive to analysis and interpretation. On incident cancer prediction, end-to-end fine-tuning achieves test AUCs of 0.81/0.75/0.73 for colorectal cancer (CRC) and 0.94/0.87/0.84 for prostate cancer across 30/60/90-day horizons on balanced cohorts, outperforming logistic-regression baselines, including time-decayed bag-of-codes. HealthFormer provides an event-centric, time-aware representation that transfers via standard fine-tuning without endpoint-specific designs. Using ICD-10 diagnoses and ATC codes can facilitate adoption beyond Hungary. Learned diagnosis embeddings align with the hierarchy, enabling clinical inspection. Broader benchmarking across endpoints remains needed.

## 1 Introduction

Longitudinal electronic health records (EHRs) provide a detailed view of patient care across time and settings. In this work, we study large-scale, patient-linked anonymized health records from the Hungarian national system^1^. These data contain sparse, irregularly timed sequences of heterogeneous clinical events spanning primary care contacts, secondary care outpatient visits, inpatient episodes, procedures, and medication dispensations. A key methodological challenge is that each event may bundle multiple co-occurring codes drawn from distinct coding systems, while inter-event gaps can range from days to years. Effective modeling therefore requires capturing both intra-event structure (code co-occurrence and cross-domain interactions) and inter-event temporal dynamics.

Sequential deep learning methods have increasingly replaced hand-crafted aggregation for EHR prediction. Early recurrent approaches demonstrated the value of temporal modeling for forecasting and interpretability [2]. More recently, Transformer-based models for EHR have leveraged masked pretraining to learn transferable representations from large-scale unlabeled records [3–5]. Despite strong empirical performance, two issues remain central for administrative and multi-domain settings: (i) flattening complex encounters into a single token or unordered bag can erase within-encounter structure, and (ii) coarse positional or bucketed time representations can underuse clinically informative time gaps [6].

We present HealthFormer, a dual-level, time-aware pretraining framework that addresses these challenges with an explicit event-centric representation and a dual-level Transformer architecture.^2^ HealthFormer first encodes the composition of an individual event using an *Intra-Event Encoder*, and then models the longitudinal patient trajectory using an *Inter-Event Encoder* equipped with an ALiBI-style continuous-time attention bias [7] and a shared date encoder. This separation preserves event-level heterogeneity while scaling to long patient histories. The same pretrained model can be adapted to new endpoints via standard fine-tuning, without designing endpoint-specific model architectures or feature sets.

### The main contributions are

- *Event-centric representation for heterogeneous administrative EHR*. We construct a unified patient timeline as an ordered sequence of typed events, each carrying a domain-specific set of codes and optional context tokens.
- *Dual-level, time-aware Transformer architecture*. We separate within-event encoding from across-event sequence modeling, and inject temporal information via a date encoder and continuous-time ALiBI bias.
- *Multi-task self-supervised pretraining*. We pretrain with complementary objectives at the code, event-type, and time-to-next-event levels, consistent with the architecture in Fig. 1.
- *Empirical analysis and downstream evaluation*. We analyze the geometry of learned ICD-10^3^ embeddings and evaluate transfer to incident cancer prediction for colorectal cancer and prostate cancer at 30/60/90-day horizons, where end-to-end fine-tuning achieves test AUCs of 0.81/0.75/0.73 for CRC and 0.94/0.87/0.84 for prostate cancer on balanced cohorts (Table 2).

**Figure 1.**
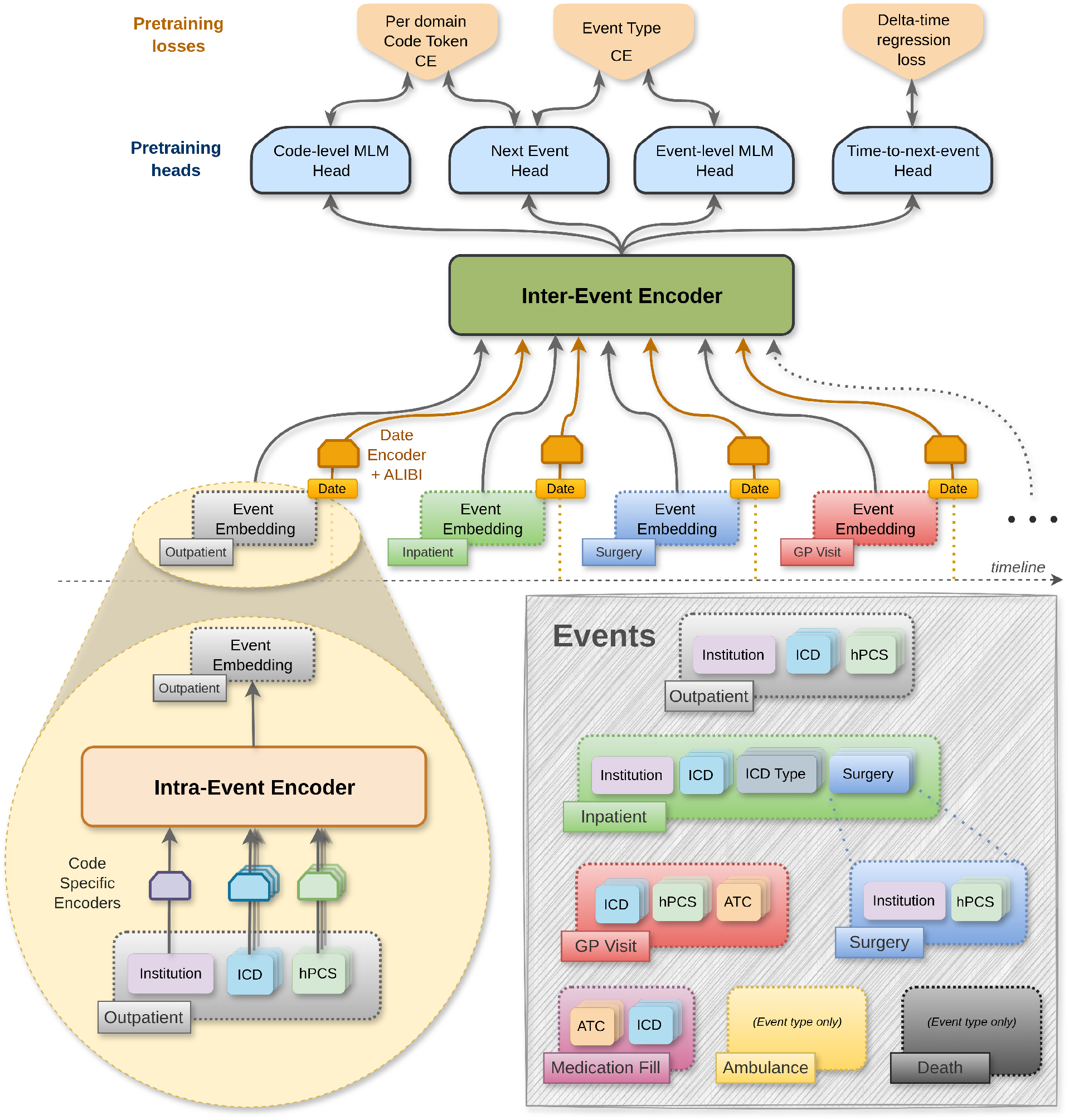
Overview of the HealthFormer architecture and pretraining objectives. Clinical records are represented as a time-ordered sequence of typed events. Within each event, heterogeneous domain tokens (e.g., ICD diagnoses, hPCS procedures, ATC medications, institution metadata) are encoded by code-specific embedding modules and aggregated by an *Intra-Event Encoder* into an event embedding. Event embeddings are combined with a shared *Date Encoder* and processed by an *Inter-Event Encoder* (Transformer) that incorporates ALiBI-based continuous-time attention biases. The pretrained sequence representation is optimized with four heads: masked code-token prediction (Code-level MLM; per-domain cross entropy), event-type prediction under full-event masking (Code-level MLM; per-domain cross entropy and Event-level MLM; cross entropy), next-event type prediction (Event-level MLM; cross entropy), and time-to-next-event prediction (Delta-time; regression loss).

## 1.1 Related work

### 1.1.1 Representation learning for structured EHR

Early deep learning approaches often serialize the patient record as a sequence of coded events and apply recurrent models for forecasting diagnoses, procedures, or outcomes. Doctor AI [1] models timestamped code streams, while RETAIN [2] adds reverse-time attention for interpretability. Other inductive biases include representation learning with denoising autoencoders (Deep Patient [8]) and convolutional architectures over embedded codes (Deepr [9]). To better capture within-encounter composition, MiME [10] models interactions between diagnoses and related interventions when multiple code types co-occur.

A recurring limitation is that many methods rely on a single “visit” abstraction that either aggregates away important care-setting and episode structure or becomes unwieldy when tokenized more finely. HealthFormer addresses this by representing the record as a sequence of typed administrative events (e.g., GP, outpatient, inpatient, surgery, medication fills), each carrying domain-specific code groups and context, and by decomposing complex episodes into atomic events to preserve clinically meaningful structure without collapsing heterogeneous encounters into one flat code bag.

### 1.1.2 Transformers and self-supervised pretraining on EHR

Transformer sequence modeling [11] and masked language modeling [12] motivated widespread EHR pretraining. BEHRT [3] adapts BERT-style objectives to longitudinal diagnosis sequences; Med-BERT [4] and TransforMEHR [5] further develop tokenization and pretraining for clinical codes and tasks. Hierarchical variants such as HiBEHRT [13] model within-visit structure, and surveys emphasize that tokenization and objectives jointly determine what clinical semantics can be recovered [14, 15].

In contrast, HealthFormer is designed as a general-purpose pretrained backbone whose representations are intended to transfer across multiple downstream clinical prediction tasks via fine-tuning. It explicitly separates (i) intra-encounter encoding from (ii) inter-encounter trajectory modeling, and trains with complementary self-supervised signals beyond standard MLM to support robust downstream adaptation.

### 1.1.3 Temporal modeling under irregular sampling

Irregular time gaps are intrinsic to care trajectories and motivate continuous-time parameterizations. Time2Vec [16] provides an expressive basis for elapsed-time embeddings, while ALiBI [7] introduces distance-dependent attention biases that can improve length generalization. In EHR modeling, time-aware attention appears in hierarchical architectures such as HiTANet [17], and self-supervised frameworks such as RAPT [6] leverage temporal structure during pretraining.

In contrast, HealthFormer injects time gaps directly into attention via a continuous ALiBI-style bias with log-scaled and clipped elapsed-time, enabling stable multi-scale temporal sensitivity and supporting both relative gaps and absolute calendar-time signals without relying on coarse time buckets.

## 2 Data representation and event construction

### 2.1 Heterogeneous data sources

We work with longitudinal, patient-linked, anonymized administrative health records from the Hungarian national system. ^4^ The dataset covers roughly 10 million individuals over a 12-year observation window. It captures time-stamped events and structured codes across a broad range of care settings, but does not include unstructured outpatient visit notes, discharge summaries, or laboratory measurements. For each individual, the data provide (i) static attributes (e.g., year of birth and biological sex) and (ii) multiple time-stamped clinical record streams originating from distinct care settings. We normalize these streams into a single event sequence representation to enable uniform modeling while retaining care-setting provenance.

Our event construction pipeline aggregates information from seven sources, each mapped to one event type (Table 1):

**Table 1:** Event taxonomy and modular composition. Each event type supports a specific subset of clinical domains. (• denotes supported, − denotes not applicable).

| Event Type ($y_t$) | Diagnosis<br>(ICD) | Procedure<br>(hPCS) | Medication<br>(ATC) | Facility<br>(Context) |
| --- | --- | --- | --- | --- |
| GP VISIT | • | — | • | — |
| OUTPATIENT | • | • | — | • |
| INPATIENT | • | — | — | • |
| SURGERY | — | • | — | • |
| MEDICATION FILL | — | — | • | — |
| AMBULANCE | — | — | — | — |
| DEATH | — | — | — | — |

**Table 2:** Test AUC for the two downstream cancer-prediction tasks across 30-, 60-, and 90-day horizons. For each method, the first row reports mean AUC and the second row reports the corresponding standard deviation over three random seeds on balanced test splits. CRC cohorts are filtered to individuals aged 50–70 years at prediction time, whereas prostate cohorts are restricted to males aged 50–70 years. To prevent label leakage, histories are truncated at a horizon-specific prediction cutoff: for positives, the cutoff is *h* days before the first diagnosis date; for negatives, matched horizon-specific cutoffs are used. Best AUC within each task-horizon pair is shown in bold.

|  | CRC |  |  | Prostate |  |  |  |
| --- | --- | --- | --- | --- | --- | --- | --- |
|  | h-30 | h-60 | h-90 | h-30 | h-60 | h-90 |  |
| *age + sex* | 0.65<br>0 | 0.65<br>0 | 0.65<br>0 | 0.71<br>0 | 0.71<br>0 | 0.71<br>0 | *AUC*<br>*Std* |
| *age + sex + util* | 0.65<br>0 | 0.65<br>0 | 0.64<br>0 | 0.79<br>0 | 0.76<br>0 | 0.75<br>0 | *AUC*<br>*Std* |
| *bag-of-codes* | 0.62<br>1.8e-5 | 0.58<br>1.1e-5 | 0.57<br>1.2e-5 | 0.79<br>1.7e-5 | 0.72<br>1.1e-5 | 0.70<br>1.6e-5 | *AUC*<br>*Std* |
| *t-decay bag-of-codes* | 0.68<br>8.5e-6 | 0.62<br>3.1e-5 | 0.60<br>2.5e-5 | 0.85<br>9.5e-6 | 0.77<br>1.9e-5 | 0.73<br>2.4e-5 | *AUC*<br>*Std* |
| *frozen HealthFormer* | 0.67<br>6.8e-3 | 0.66<br>3.0e-3 | 0.66<br>1.4e-3 | 0.82<br>2.3e-3 | 0.78<br>5.7e-4 | 0.77<br>2.6e-3 | *AUC*<br>*Std* |
| *finetuned HealthFormer* | **0.81**<br>**1.5e-3** | **0.75**<br>**2.9e-3** | **0.73**<br>**3.1e-3** | **0.94**<br>**9.4e-4** | **0.87**<br>**1.4e-3** | **0.84**<br>**1.8e-3** | *AUC*<br>*Std* |

#### GP encounters

Primary care contacts containing diagnoses and prescribed medications (GP Visit).

#### Secondary and tertiary care outpatient visits

Ambulatory specialty care records with diagnoses and procedures, together with facility context (Outpatient).

#### Inpatient episodes

Hospital admissions with admission/discharge intervals and discharge diagnoses, with facility identifiers (Inpatient).

#### Surgeries and interventions

Procedure-centric records anchored to intervention dates, typically with facility context (Surgery).

#### Medication dispensations

Prescription fill records linking individuals to medication products/classes (Medication Fill).

#### Ambulance transport

Ambulance transport records, treated as a distinct care-setting signal even when structured code payloads are sparse (Ambulance).

#### Death

When available in the administrative record, a death date is represented as a terminal event without structured code payload (Death).

These streams differ in temporal semantics: inpatient episodes span multi-day intervals, whereas most other records are point events. Timestamp completeness and precision also vary by source. Before sequence construction, we harmonize dates, remove records with missing or sentinel timestamps, and resolve duplicates deterministically.

### 2.2 Unified event taxonomy

We represent a patient’s history as a sequence ℰ= [*e*_1_, *e*_2_, …, *e*_*T*_]. Each event *e*_*t*_ is a typed container

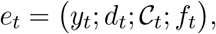

where 𝒴_*t*_ is the event type, *d*_*t*_ is the event date, 𝒞 _*t*_ is a set of domain-specific code groups, and *f*_*t*_ contains optional context metadata (e.g., institution identifiers).

Clinical codes originate from multiple coding systems. We use ICD for diagnoses, hPCS for procedures, and medication information via ATC classes. Some metadata (e.g., institution) are treated as additional tokens. Event types support different combinations of domains (Table 1); for example, a Surgery event may include procedure codes and facility context, while Ambulance and Death events may be represented by the event type alone.

#### Decomposition of complex episodes

Certain raw records are decomposed into multiple atomic events to preserve temporal fidelity. For example, an inpatient case is represented as an Inpatient event anchored at admission, while surgeries recorded during the stay are emitted as distinct Surgery events anchored to their intervention dates. This design separates episode context from specific interventions.

#### Deterministic sequence construction

To produce reproducible timelines, we aggregate all events for a patient and sort them by date. When multiple events share the same day, we break ties using a fixed precedence order designed to reflect typical clinical workflow.

### 2.3 Temporal signals

Standard positional encodings are ill-suited for irregular sampling. HealthFormer derives time signals from event dates and injects them through (i) a date encoder at the event level and (ii) a continuous-time attention bias at the sequence level. In addition to absolute date (in days), we derive inter-event gaps (Δ*t*) and patient age at the event time when available; for events nested within an inpatient interval, we also compute an episode-relative offset. Implementation details of time-feature computation and the exact discretization bin edges are reported in Appendix B (Table B.1).

### 2.4 Hierarchical tokenization

We map structured clinical codes to learnable token IDs with domain-specific tokenization.

#### Hierarchical expansion (ICD and ATC)

Diagnoses (ICD) and medication classes (ATC) are defined by hierarchies. We apply hierarchical expansion by decomposing each code into its ancestor path up to a configured depth. Each expanded node is represented by the sum of a node embedding and a learned depth embedding, allowing statistical sharing across related codes.

#### Flat and hashed vocabularies

Procedure codes (hPCS) are mapped to flat vocabularies. High-cardinality metadata (e.g., institution identifiers) are mapped using deterministic feature hashing into a fixed number of buckets to bound memory while retaining site/context signal.

#### Token budgets

Events vary widely in complexity. We enforce reasonably large per-domain token budgets (e.g. diagnoses and procedures) and deterministically truncate events that exceed these limits. Complete tokenization hyperparameters (domain budgets, hierarchy depth, hashing) are provided in Appendix A, Table A.1.

## 3 HealthFormer architecture

HealthFormer is a dual-level Transformer designed for irregular sequences of heterogeneous clinical events. It separates (i) within-event encoding of multi-domain tokens from (ii) across-event sequence modeling, and integrates time through a shared date encoder and ALiBI-based attention bias (Fig. 1).

### 3.1 Notation

For a patient, let 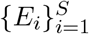 denote a sequence of *S* events. Event *E*_*i*_ contains: (i) an event type *e*_*i*_ ∈{1, …, *E*}, (ii) an event time *t*_*i*_ (days), and (iii) a variable-length set of tokens 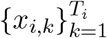 with domain labels *d*_*i,k*_. We use model width *d* and *H* attention heads, with padding masks for tokens within an event 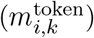 and events within a patient 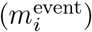.

### 3.2 Code-specific embedding modules

Clinical events combine multiple code systems and metadata fields, each with different vocabulary sizes and semantics. We therefore use code-specific embedding modules per domain and project all token vectors into a shared *d*-dimensional space before attention-based aggregation.

#### Flat code embeddings

For domains treated as atomic symbols, each token receives a domain-specific embedding

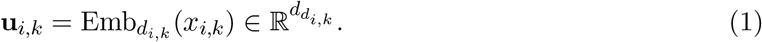

#### Hierarchical code embeddings

For hierarchical domains (e.g., ICD, ATC), we combine node and depth embeddings. Let depth(*x*_*i,k*_) ∈{0, 1, …} denote a depth index; then

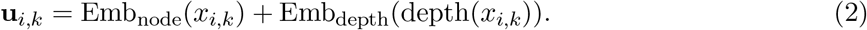

#### Projection to a shared model space

Because domains may use different embedding widths *d*_*di,k*_, we project to a common model width *d*:

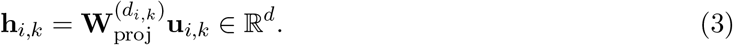

After padding and concatenating all token groups within *E*_*i*_, we obtain the per-event token matrix **H**_*i*_ ∈ ℝ^*T* ×*d*^.

### 3.3 Intra-Event Encoder

The Intra-Event Encoder aggregates the unordered set of within-event tokens into a single event embedding. We implement attention pooling so that the model can weight tokens differently depending on the event context. Concretely, the query is derived from an event-type embedding, while keys and values come from within-event token embeddings.

#### Query, keys, and values

Let **e**_*i*_ = Emb_type_(*e*_*i*_) ∈ ℝ^*d*^. We compute

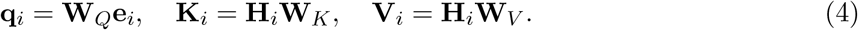

#### Masked attention pooling

For head *h* and token position *k*, attention weights are

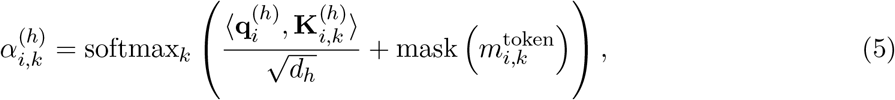

with pooled output

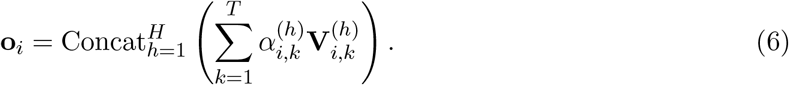

We add a residual skip connection from an event-type prior **s**(*e*_*i*_):

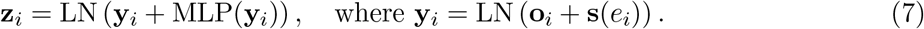

### 3.4 Date encoder and time integration

To encode absolute time, we compute a time embedding 𝒯_*i*_ from the event date (and derived signals such as age) using a sinusoidal Time2Vec basis [16] and inject it additively:

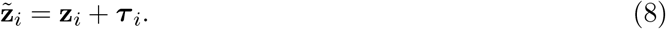

This provides each event representation with an explicit time context; relative time effects are handled by the attention bias described below.

### 3.5 Inter-Event Encoder with continuous-time ALiBI

We model longitudinal progression using a Transformer encoder stack over the event sequence. To capture the continuous nature of irregular time gaps, we implement a log-scaled version of ALiBI, where the attention bias is derived from the logarithm of the elapsed time rather than token position.

#### Time-aware attention bias

Define the time gap Δ_*i,j*_ = |*t*_*i*_ − *t*_*j*_|. For each head *h*, we add a fixed bias to attention logits:

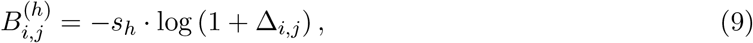

where *s*_*h*_ is a head-specific slope from a fixed geometric sequence. This bias encourages attention locality in physical time while allowing long-range interactions.

#### Transformer layer

Attention logits are

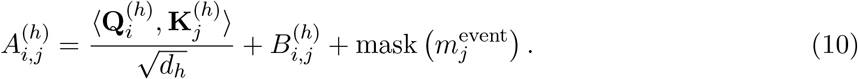

Stacking layers yields contextualized event representations 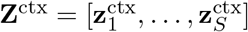.

### 3.6 Patient-level pooling

For downstream tasks, we map the contextualized event sequence 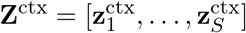 to a fixed-size patient representation using learned single-query attention pooling over valid events. For each event *i*, we compute

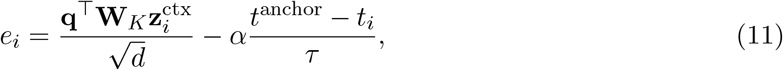

where **q** is a learned query vector, **W**_*K*_ is a learned projection, *t*^anchor^ is the most recent event time in the sequence, and the second term is a learnable recency bias. These scores are softmax-normalized over valid events to obtain attention weights, and the final patient representation is

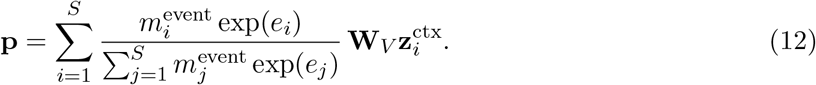

All encoder architectural hyperparameters are summarized in Appendix C (Table C.1).

## 4 Self-supervised pretraining

HealthFormer is pretrained using multiple self-supervised objectives defined on the same event-centric representation. The overall training signal is a weighted sum of losses from four heads (Fig. 1):

### 1. Code-level MLM (masked code-token prediction)

1. We randomly mask code tokens and train the model to recover them from (i) the remaining codes in the same event and (ii) longitudinal context. Because domains have distinct vocabularies, masked-token prediction is implemented with a *per-domain* classification head and cross-entropy loss (“per-domain Code Token CE”).

### 2. Event-level MLM (event-type inference under full-event masking)

With a fixed probability, we mask an entire event (and its belonging codes) by replacing its event type with a dedicated mask token and removing its code payload. The model must infer the original event type using surrounding events. We supervise with cross-entropy over event types.

### 3. Next-event prediction (event type and codes)

For each position *t*, we use the representation at *t* to predict the *next* event at *t*+1, including both its event type and its code tokens. Supervision is defined by a one-step shift of the original (unmasked) sequence, with cross-entropy losses over event types and per-domain code vocabularies. To prevent information leakage, this head is computed under a *strict causal inter-event attention mask* : when forming the representation at *t*, attention to all positions > *t* is blocked, so no information from the target event at *t*+1 (neither type nor codes) is visible to the model.

### 4. Time-to-next-event (Δ*t* prediction)

For each position *t*, we predict the elapsed time in days until the next event,

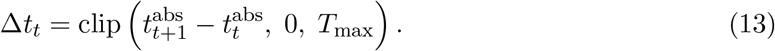

supervised with a regression loss (we use *L*_1_ loss on the clipped gap; “delta-time regression loss”). Objective-specific masking/targeting details and loss weights are given in Appendix D (Table D.1).

## 5 Supervised finetuning

To assess transfer to supervised endpoints, we attach a lightweight classification head to the pretrained HealthFormer encoder and optimize a standard supervised objective on labeled patient histories. Because HealthFormer operates on a generic event-centric representation with shared tokenization and time-aware sequence modeling, the same pretrained backbone can be adapted to a wide range of downstream problems (e.g., incident diagnosis prediction, risk stratification, utilization forecasting, or other supervised endpoints) without task-specific architectural changes.

### Adaptation regimes

We consider two practical transfer regimes. In *end-to-end fine-tuning*, all encoder parameters and the task head are optimized jointly, enabling endpoint-specific adaptation of representations. In *frozen-encoder* (probe-style) training, the pretrained encoder is kept fixed and only the task head is learned, isolating the usefulness of pretrained representations for a given task. Experimental results are described in the corresponding downstream Section 6.2.

## 6 Results

### 6.1 Intrinsic structure of ICD embeddings

To qualitatively assess whether self-supervised pretraining induces clinically meaningful geometry in the learned diagnosis space, we visualize the ICD token embeddings learned under the *Code-level MLM* objective.^5^ Because Code-level MLM operates directly at the code level, it provides a strong training signal to the token embedding pathway (in addition to the higher-level sequence model), making it a natural probe of the intrinsic organization of the learned code space.

#### Embedding extraction and representation

We extract the ICD embedding lookup table from the pretrained checkpoint and focus on hierarchy depth 2 (the finest granularity used in our runs with depths 0/1/2). Each marker corresponds to one depth-2 ICD node represented by its *learned lookup embedding* (72 dimensions), i.e., the vector stored in the embedding table before any encoder-side transformations. During encoding, hierarchical tokenization also adds a learned depth embedding; here we omit depth embeddings to isolate structure learned in the lookup table itself rather than the depth index. Reserved indices (padding/mask) are excluded.

#### Dimensionality reduction and visualization

We apply PCA, UMAP, and t-SNE to the extracted vectors, obtaining qualitatively consistent patterns across methods. We report t-SNE in Fig. 2 (perplexity 30, learning rate 200) to highlight local neighborhood structure. Points are colored by the parent depth-0 category and sized by empirical code frequency in the pretraining corpus.

**Figure 2.**
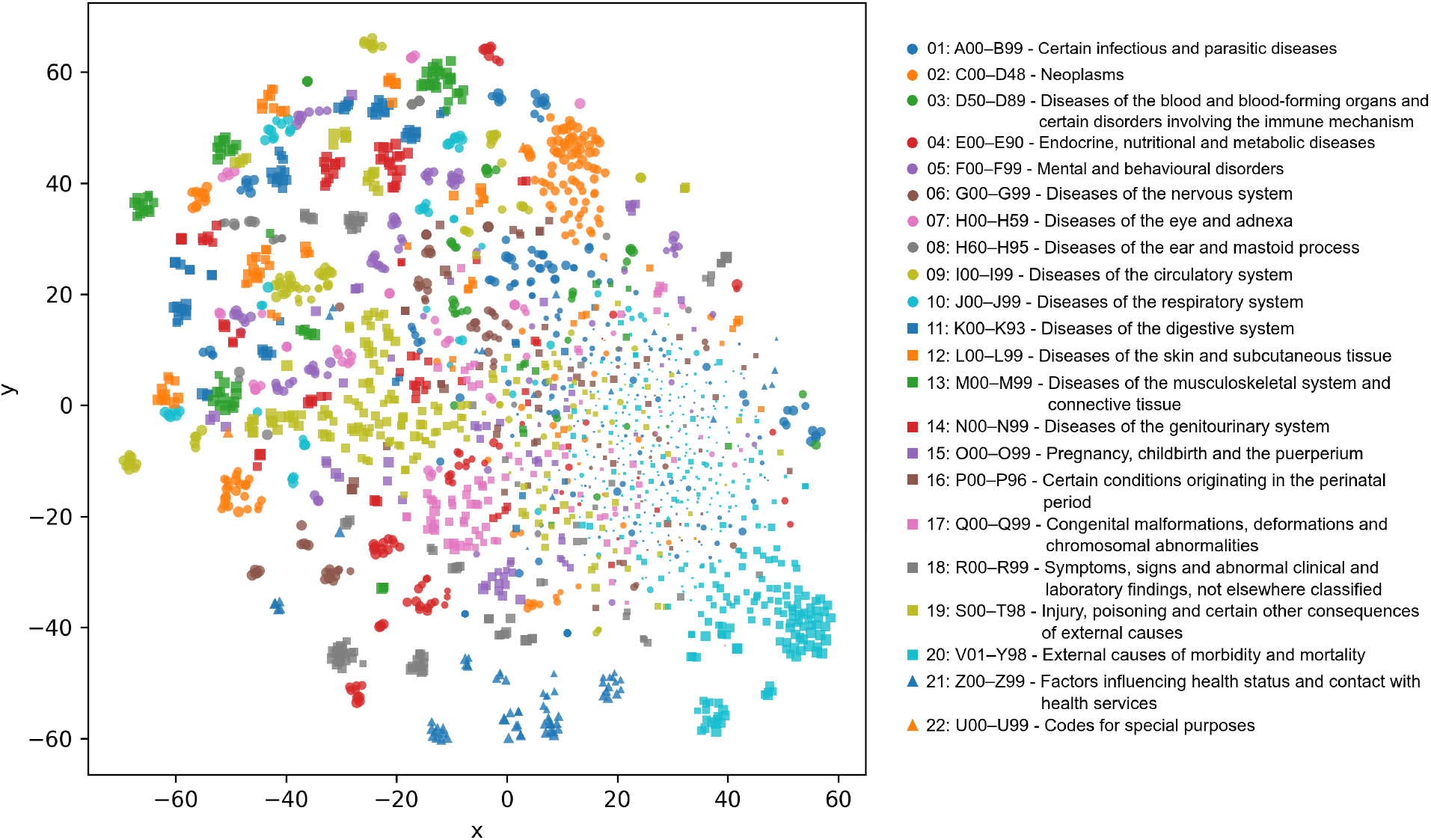
t-SNE visualization of depth-2 ICD lookup embeddings learned during MLM pretraining. Each point is a depth-2 ICD code embedding (72-dimensional vector from the embedding table). Colors indicate the corresponding depth-0 category, and marker size reflects empirical code frequency in the pretraining data.

#### Qualitative findings and interpretation

The map exhibits clear local clustering: within many depth-1 groups, depth-2 codes form compact neighborhoods, indicating coherent organization of fine-grained diagnoses in the learned space. In contrast, broader depth-0 aggregates do not necessarily form a single contiguous region; their constituent depth-1 clusters can be scattered (note that Fig. 2 uses depth-0 coloring), and even within a single color we observe multiple spatially separated sub-clusters; these sub-clusters align closely with depth-1 groupings (details in Section F). Two factors could promote hierarchy-aligned structure: (i) the model has access to depth information via an explicit depth embedding during encoding, and (ii) Code-level MLM couples codes through shared intra-event and longitudinal context, drawing frequently co-occurring or substitutable codes closer. However, because Fig. 2 visualizes only the depth-2 *lookup* embeddings (excluding depth embeddings), the observed organization cannot be attributed solely to injecting the depth index, suggesting that pretraining shapes the embedding table itself.

We also observe a frequency-dependent pattern: along the periphery of the map, several categories form well-separated clusters, whereas the lower-right quadrant contains more blended, overlapping regions. Notably, the well-clustered areas tend to contain larger markers, while mixed/noisy regions are dominated by smaller markers. Since marker size reflects empirical code frequency, this suggests that the method produces more stable, cluster-consistent embeddings for data-rich ICD codes, while less frequent codes are embedded in a noisier, less separable manner.

Beyond hierarchy-consistent clustering, we observe early qualitative signals that some neighborhoods align with clinically plausible co-occurrence relationships (e.g., symptom constellations, common differentials, or shared etiologies). These effects are not uniform and should be interpreted cautiously; we leave systematic validation to future work, including quantitative neighborhood analyses and downstream fine-tuning.

### 6.2 Incident cancer prediction

We evaluate HealthFormer on two incident cancer prediction tasks: colorectal cancer (`crc`) and prostate cancer (`prostate`) under three prediction horizons: 30, 60, and 90 days. For each horizon *h* ∈{30, 60, 90}, we construct an input history ending at a prediction cutoff and predict whether a first diagnosis occurs within the subsequent *h*-day window. Across both tasks, we reuse the same pretrained encoder, tokenizer, and downstream pooling mechanism, preserving a common event-centric transfer pipeline.

#### Cohort construction and leakage control

All cohorts are filtered to include individuals aged 50–70 years at prediction time; prostate cohorts are additionally restricted to males. For positive individuals, we truncate the observed history to end exactly *h* days before the first diagnosis date to avoid label leakage. For each task and horizon, we construct balanced patient-level splits with 15,000 positives and 15,000 negatives for training and 1,000 positives and 1,000 negatives each for validation and test sets. We report area under the receiver operating characteristic curve (*AUC*) on test splits, averaged over three random seeds. Additional details on task-specific cohort construction and case definitions are provided in Appendix E.1.

#### Model regimes and baselines

We compare HealthFormer under two supervised adaptation regimes (end-to-end fine-tuning and frozen-encoder probing; Section 5) and several lightweight logistic-regression baselines. Hyperparameters are reported in Appendix E.2 (Table E.1).

`Age + sex` *(logistic regression)*. Uses age (years) at cutoff time and a one-hot encoding of sex.

`Age + sex + util` *(logistic regression)*. Extends age+sex with coarse utilization summaries computed on the truncated history: total number of events, counts by event type, time since last event (days), and total history span (days).

`Bag-of-codes` *(logistic regression)*. Uses a sparse count vector over observed structured codes. For each event, we add +1 to features of the form `domain:code` for codes appearing in the event (ICD, hPCS, ATC, and facility identifiers when available). To mirror the model-side input constraints, we (i) keep only the most recent events up to the same maximum event-sequence length and (ii) apply the same per-domain token limits within each event.

`Time-decay bag-of-codes` *(logistic regression)*. Augments bag-of-codes with an exponential recency weight. For an event occurring *δ* days before cutoff time, the feature increment is multiplied by

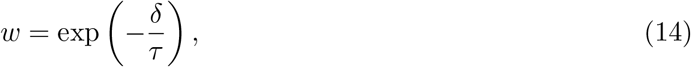

with 𝒯 = 365 days.

Additional details on baseline feature construction and logistic regression solver settings are provided in Appendix E.3.

#### Results summary

Table 2 reports test AUC across both cancer-prediction tasks and all three horizons. End-to-end fine-tuning yields the strongest performance in every reported setting, reaching AUCs of 0.81/0.75/0.73 for CRC and 0.94/0.87/0.84 for prostate prediction at 30/60/90 days, respectively. On CRC, the fine-tuned model improves substantially over the strongest logistic-regression baseline, the time-decay bag-of-codes model, by approximately 0.13 AUC at each horizon. On prostate prediction, the gains are similarly consistent, with margins of 0.09, 0.10, and 0.11 AUC over the same baseline at 30, 60, and 90 days. These results indicate that the pretrained event-centric sequence model captures temporally structured signal beyond what can be recovered from recency-weighted counting alone, and that this advantage transfers across distinct incident-cancer endpoints.

A practical advantage of the HealthFormer pipeline is that the same pretrained encoder and tokenizer can be reused across endpoints while preserving a common event-centric representation. This enables efficient adaptation to new supervised tasks without redesigning the input representation or constructing endpoint-specific deep architectures.

## 7 Discussion

HealthFormer is motivated by two practical properties of longitudinal administrative EHR: heterogeneous event composition and highly irregular sampling. By explicitly separating intra-event encoding from inter-event sequence modeling, the framework preserves encounter-level structure while supporting long timelines. The embedding analysis suggests that self-supervised pretraining shapes diagnosis representations in a way that is consistent with the underlying ICD hierarchy, revealing clinically interpretable neighborhoods in the learned code space.

In downstream evaluation, HealthFormer improves over logistic-regression baselines that incorporate demographic information, utilization summaries, and time-decayed bag-of-codes features. While these baselines are lightweight, they are commonly used in administrative settings and provide a meaningful reference point for assessing whether contextual sequence modeling adds value beyond recency-weighted counting. An additional practical strength of the approach is adaptation: the same pretrained encoder can be fine-tuned end-to-end for a new endpoint without constructing a task-specific architecture or feature representation.

### Limitations and future work

This study has several limitations and clear opportunities for extension. Downstream evaluation currently covers two incident cancer endpoints (CRC and prostate cancer) and mainly compares against logistic-regression baselines; a natural next step is a broader benchmark across additional endpoints and external validation, including comparisons to strong deep sequential models trained on the same event-centric representation. The event-centric design enables clinically useful interpretability analyses: by attributing predictions to specific prior events and code groups, we can identify which historical signals most strongly support a given endpoint. This opens the door to systematically mapping which outcomes are predictably driven by which patterns in patient trajectories, helping prioritize candidate endpoints, refine feature sets, and generate testable hypotheses for follow-up studies. Beyond structured administrative code streams, the architecture is designed to incorporate additional modalities, such as numeric laboratory results and richer clinical documentation, which may further improve clinical utility. Finally, deployment-oriented work should include calibration, robustness, and bias analyses to support safe use in practice.

## Conclusions

HealthFormer provides a dual-level, time-aware pretraining framework for irregular EHR event sequences. Its event-centric representation and multi-task self-supervision yield embeddings with interpretable structure and enable practical transfer via standard fine-tuning.

## Acknowledgements and Declarations

## Acknowledgements

The authors acknowledge the data custodian and collaborators who enabled access to the Hungarian administrative health records.

## Funding

This work was supported by the National Research, Development and Innovation Office (NKFIH) within the framework of the Artificial Intelligence National Laboratory under project identifier RRF-2.3.1-21-2022-00004, and by the National Research, Development and Innovation Office (NKFIH) within the framework of the National Laboratory for Health Security under project identifier RRF-2.3.1-21-2022-00006.

## Competing interests

The authors declare that they have no known competing financial interests or personal relationships that could have appeared to influence the work.

## Data availability

The Hungarian administrative health records used in this study are subject to legal and governance restrictions and are not publicly available. Derived datasets supporting the findings of this study can be made available from the authors upon reasonable request and with permission of the data custodian.

## Ethics approval and informed consent

We conducted a retrospective cohort study using depersonalised administrative health (for billing purposes) data from the Hungarian National Health Insurance Fund (NHIF) database, available for the period January 2010 to December 2021. The database was depersonalised by replacing real-world personal identification codes with generated identifiers. In accordance with the European General Data Protection Regulation (GDPR) and national regulations, ethical approval for the study was obtained (License No.: BM/14830-3/2024). Informed consent was waived because this was a retrospective study using depersonalised administrative data, in accordance with applicable national regulations.

## Declaration of generative AI and AI-assisted technologies in the manuscript preparation process

During the preparation of this work, the authors used ChatGPT (OpenAI) to improve the clarity and readability of the manuscript. After using this tool, the authors reviewed and edited the content as needed and take full responsibility for the content of the published article.

## Appendix: Implementation Details and Additional Results A

### Event construction, ordering, and tokenization

#### A.1 Event types and deterministic ordering

Each patient record is represented as a time-ordered sequence of typed events. Events are sorted primarily by calendar date; ties on the same day are broken deterministically using a fixed type-priority order and then by remaining fields (codes and metadata), ensuring reproducible serialization.

##### Type priority for same-day ties

When multiple events share the same day, the following precedence is used (highest priority first): D<sc>eath</sc> > I<sc>npatient</sc> > S<sc>urgery</sc> > O<sc>utpatient</sc> > GP<sc> visit</sc> > P<sc>rescription fill</sc> > A<sc>mbulance</sc>.

#### A.2 Token serialization and reserved IDs

##### Reserved token IDs

Across all code domains and event types, two IDs are reserved:

- **0**: padding (`PAD`)
- **1**: mask token (`MASK`)

All non-reserved (real) tokens therefore start at ID 2. For event types, the mapping is implemented as `event_type_id = 2 + index(EventType)`; the event-type masking objective uses MASK (ID 1).

##### Sequence truncation

The maximum event-sequence length is `max_seq_len` = 512. With `truncate_direction = right`, sequences longer than 512 events are truncated to the *most recent* events, and shorter sequences are left-padded to length 512 (padding occurs at the beginning of the sequence).

#### A.3 Domain budgets, hierarchical expansion, and hashing

Within each event, the model operates on a fixed-size concatenation of per-domain token slots. The concatenation order is:

<Preformate>

[event type] ∥ [ICD] ∥ [hPCS] ∥ [ATC] ∥ [facility].

*</preformate*>

Slots not filled by real tokens are padded (ID 0) and excluded by a domain-specific token mask.

##### Hierarchical expansion (ICD, ATC)

For ICD and ATC, each code is expanded into ancestor nodes up to a maximum depth of 2 (`hierarchical_depth=2, hierarchical_min_depth=0`). Each expanded node token is represented by the sum of (i) a node embedding and (ii) a learned depth embedding. When the expanded node list exceeds the per-event token budget, the remainder is truncated.

**Table A.1:** Tokenizer and per-domain budgets from `config.yaml`. The total token slots per event are 1 + 50 + 25 + 0 + 10 + 1 = 87. For hierarchical domains (ICD, ATC), the budget counts *expanded hierarchy nodes* rather than raw codes.

| Domain | Enabled | Max tokens/event | Hier. depth | Emb. dim (pre-proj) |
| --- | --- | --- | --- | --- |
| Event type | yes | 1 | – | 48 |
| ICD (diagnosis) | yes | 50 | 2 | 72 |
| hPCS (procedure) | yes | 25 | – | 48 |
| ATC (drug class) | yes | 10 | 2 | 36 |
| Facility (context) | yes | 1 | – | 24 |

##### Facility hashing

Facility identifiers are hashed into `facility_hash_size` = 612 buckets using SHA-1, with the resulting bucket index shifted by +2 to preserve the PAD/MASK reservation. Collisions are therefore possible by design.

**ICD-type hashing (visit modifier)**. If present, a categorical ICD modifier (`ICD_TYPE`) is hashed into `icd_type_hash_size` = 32 buckets (also shifted by +2) and added as an extra embedding component to each ICD token within the event.

### B Time features and discretization

Time features are computed per event and then aligned to the (possibly truncated) event sequence. All time quantities are represented in **days**.

#### Continuous features

For an event sequence with dates *d*_1_, …, *d*_*L*_ (in days), we compute:

1. *t*_abs_(*i*): absolute day ordinal for event *i*.
2. *t*_gap_(*i*) = clip *t*_abs_(*i*) − *t*_abs_(*i* − 1), 0, 730 for *i* > 1, and *t*_gap_(1) = 0.
3. *t*_age_(*i*): age in days at event *i* (derived from the stored birth date; first event defines the base).
4. *t*_epi_(*i*): days since the start of an inpatient episode if the event lies within an episode; otherwise a sentinel value `-1`.

#### Discretized bins (packaged defaults)

Let *E* denote the monotonically increasing edge vector; bin indices are computed as

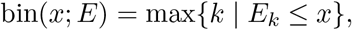

clipped to [0, |*E*| − 1]. Values with *t*_epi_ *<* 0 keep the sentinel `-1`..

**Table B.1:** Time-bin definitions.

| Feature | Discretization (days) |
| --- | --- |
| Inter-event gap ($t_{\text{gap}}$) | Log-spaced bins with base $b = 1.3$ up to $T_{\text{max}} = 1825$ days ($\sim 5$ years).<br>Bin edges are generated as<br>$E_{\text{gap}} = \left\{ \lfloor b^k \rfloor \mid k = 0, 1, \dots, K \right\} \cup \{T_{\text{max}}\}, \quad K = \max\{k : \lfloor b^k \rfloor < T_{\text{max}}\},$ followed by sorting and removing duplicates. |
| Within-episode offset ($t_{\text{epi}}$) | Fixed edges $E_{\text{epi}} = \{0, 4, 11\}$, corresponding to bins $[0, 3]$, $[4, 10]$, and $\geq 11$ days. A sentinel value of $-1$ is used for non-episode events. |

#### Calendar embeddings

Month-of-year and day-of-week are also embedded (default dimensions: 16 each) and fused into the time representation.

### C Encoder architecture hyperparameters

This section documents the exact architectural hyperparameters of the pretrained encoder.

#### C.1 High-level architecture

**Table C.1:** Encoder hyperparameters from config.yaml (and corresponding code defaults for components not explicitly parameterized).

| Component | Setting |
| --- | --- |
| Event embedding dimension ($d$) | 192 |
| Time embedding dimension ($d_{\text{time}}$) | 48 (fused into $d$) |
| Intra-event encoder | attentive pooling, 4 heads; MLP hidden dim = $d$; dropout 0.1 |
| Inter-event encoder | 5 layers; 8 attention heads; FFN dim 1024; dropout 0.1; GELU |
| Normalization | LayerNorm in intra- and inter-event blocks |
| Sex embedding | enabled; embedding dim 8 (projected to $d$) |
| Sequence pooling outputs | `pooled_last`, `pooled_mean` and `sequence_output` available |

#### C.2 ALiBi time bias parameters

The inter-event transformer incorporates ALiBi-style attention logit biases based on pairwise absolute time differences Δ*t*_*ij*_ = |*t*_abs_(*i*) − *t*_abs_(*j*)| in days. The configured time transform is:

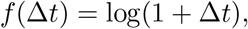

and the per-head slopes (packaged defaults) are:

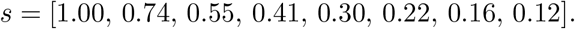

#### C.3 Vocabulary sizes

Domain vocabularies are constructed from external code tables (ICD, ATC, hPCS vocabularies), with hierarchical expansion increasing the effective node vocabulary for ICD/ATC.

### DPretraining objectives and optimization hyperparameters

#### D.1 Objective hyperparameters and weights

Pretraining optimizes a weighted sum of four objectives. The loss weights and objective-specific parameters are:

**Table D.1:** Pretraining objective configuration (from config.yaml and objective implementations).

| Objective | Hyperparameters (as implemented) |
| --- | --- |
| Code-level MLM | Mask probability 0.15 per (event, token-slot, domain) on valid code slots. 80/10/10 masking policy: 80% replaced by **MASK** (ID 1), 10% replaced by a random valid ID ($\geq 2$), 10% left unchanged. Labels use **ignore_index=0** for non-masked and padded slots. |
| Event-level MLM | Mask probability 0.15 per event on valid (non-padding) events. Masked events set event-type ID to **MASK** (ID 1) and zero-out all code tokens for that event. Labels use **PAD** (ID 0) as ignore index. |
| Next-event prediction | Predicts (a) next event type and (b) next-event codes from the current event embedding (shift by one). Both type and code losses are enabled; weights: type=1.0, code=1.0. Pairs involving padding are excluded. |
| Delta-time prediction | Mode: regression; loss: L1. Targets are $\Delta t$ between consecutive events in days, clamped to max_gap_days=365. |
| Loss weights | $\lambda_{\text{code-MLM}} = 1.0$, $\lambda_{\text{event-MLM}} = 1.0$, $\lambda_{\text{next-event}} = 1.0$, $\lambda_{\Delta t} = 0.2$. |

##### Event-level code prediction

For code prediction heads (Code-level MLM and next-event code prediction), logits are produced *per event and domain* and then broadcast across token slots within that domain. This realizes a bag-of-codes style loss in which each within-event token slot is scored against the same event-level distribution.

#### D.2 Optimization and training schedule

**Table D.2:** Pretraining optimization hyperparameters (from config.yaml and training loop defaults).

| Training setting | Value |
| --- | --- |
| Optimizer | AdamW (PyTorch defaults for $\beta$ and $\epsilon$) |
| Learning rate | $3 \times 10^{-4}$ |
| Weight decay | $10^{-2}$ |
| Gradient clipping | global norm $\leq 1.0$ |
| Warmup schedule | linear warmup for 5,000 steps, then constant LR |
| Max steps (configured) | 1,500,000 |
| Batch size | 10 |
| Max validation / test examples | 10,000 each (subsample cap) |
| Logging frequency | every 50 steps |
| Eval / checkpoint frequency | every 10,000 steps |
| DataLoader workers | 32 |

### EDownstream fine-tuning and baseline configurations

#### E.1 Downstream data collection

##### CRC data collection

CRC positives were derived by a rule-based phenotyping algorithm that assigns a first CRC diagnosis date from routinely collected administrative encounters. We searched diagnosis codes (ICD) by *prefix match* against the following prefixes: C1800--C1890, C19H0, C20H0, C2100, C2110, C2120, C2180, D0100, D0110--D0130, D3730, D3740, D3750. Evidence was taken only from inpatient and outpatient records (GP and prescription ICD evidence disabled). Inpatient evidence required ss1 qualifying admission with a CRC ICD recorded either as the *primary discharge diagnosis* or as a clinically relevant *comorbidity* (the two diagnosis-role categories retained in our data), and excluded admissions belonging to pre-specified non-eligible DRG groups (per the administrative DRG codebook used in this dataset). Outpatient evidence required ≥2 qualifying visits where the CRC ICD was recorded as the main diagnosis; outpatient visits from pre-specified excluded specialties were removed, where specialty was derived from clinic ID using the study’s specialty mapping table. After the minimum-evidence criteria were satisfied (≥1 inpatient admission and/or ≥2 outpatient visits), the CRC index date was set to the earliest date among all qualifying CRC-coded encounters.

##### Prostate data collection

Prostate positives were derived by a rule-based phenotyping algorithm that assigns a first prostate-cancer diagnosis date from routinely collected administrative encounters. We searched diagnosis codes (ICD) by *prefix match* against `C61H0.` Evidence was taken only from inpatient and outpatient records (GP and prescription ICD evidence disabled). Inpatient evidence required ≥1 qualifying admission with a prostate ICD recorded either as the *primary discharge diagnosis* or as a clinically relevant *comorbidity* (the two diagnosis-role categories retained in our data), and excluded admissions belonging to pre-specified non-eligible DRG groups (per the administrative DRG codebook used in this dataset). Outpatient evidence required ≥2 qualifying visits where the prostate ICD was recorded as the main diagnosis; outpatient visits from pre-specified excluded specialties were removed, where specialty was derived from clinic ID using the study’s specialty mapping table.

In addition, cases were retained only if corroborating treatment evidence was present. Treatment evidence was taken from inpatient and outpatient procedure records using a curated list of prostate-related procedure codes, and from prescription records using ATC prefixes `G03HA01, L02AE01--L02AE04, L02BB02--L02BB05, L02BX03, L01CD02, L01CD04,` and `L01DB07.` Procedure evidence required ≥1 qualifying inpatient record and/or ≥2 qualifying outpatient records; prescription evidence required ≥1 qualifying fill. After the diagnosis and treatment criteria were satisfied, the prostate index date was set to the earliest qualifying diagnosis-coded encounter, provided that the most recent qualifying treatment record was not more than 182 days earlier than that diagnosis date.

#### E.2 HealthFormer downstream classifier hyperparameters

Downstream prediction uses a classification head on top of a learned query-attention pooled patient representation computed from the encoder sequence output, with attention restricted to valid events and augmented by a learnable recency bias.

Two training regimes are supported: (i) *freeze* (encoder frozen, head trained), and (ii) *finetune* (encoder and head trained).

**Table E.1:** Default downstream hyperparameters from packaged downstream configs.

| Setting | Value (downstream config defaults) |
| --- | --- |
| Pooling | query_attention |
| Head type | MLP |
| Head hidden dim | 128 |
| Head dropout | 0.2 |
| Batch size | 32 |
| Optimizer | AdamW with separate parameter groups (encoder vs head) |
| Encoder LR | $1 \times 10^{-5}$ |
| Head LR | $1 \times 10^{-4}$ |
| Encoder weight decay | 0.0 |
| Head weight decay | $1 \times 10^{-3}$ |
| Epochs | 1 |
| Evaluation frequency | every 4 epochs (if training for multiple epochs) |

#### E.3 Logistic regression baselines: features and solver

All baselines are trained with scikit-learn logistic regression using:

- `max_iter`=500,
- solver `saga` for sparse feature matrices (bag-of-codes) and `lbfgs` for dense features,
- default L2 regularization strength (C=1.0) unless overridden.

##### Feature sets

The baseline feature families referenced in the main text are implemented as follows:

- **Age+Sex:** dense vector (age years, ⊮[male], ⊮[female]).
- **Age+Sex+Utilization:** Age+Sex plus total number of events, per-event-type counts, time since last event, and overall history span (days).
- **Bag-of-codes:** sparse counts over observed structured codes using features of the form domain:code. To mirror model-side constraints, this baseline uses the same maximum event-sequence length (512) and per-event token budgets (Table A.1).
- **Time-decay bag-of-codes:** like bag-of-codes, but each code occurrence at event time *t* receives weight exp 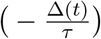with Δ(*t*) = days between event date and the cutoff date and *τ* = 365 days (default).

### F Cross-category neighborhoods in the ICD embedding map

To assess whether the learned code space captures clinically meaningful relations beyond the explicit ICD hierarchy, we inspected local neighborhoods in the t-SNE projection of depth-2 ICD lookup embeddings. While most clusters follow the taxonomy, we also observe recurring *cross-category* proximities, where codes from different depth-0 groups appear as near neighbors. These patterns are plausibly driven by shared clinical context and co-recording in longitudinal health records.

#### (1) Malignancies with mycoses and glaucoma

Malignancy-related codes often appear near fungal infections, consistent with susceptibility to opportunistic infections (such as mycosis) due to therapy induced immunosuppression; conversely, fungal infections (especially candidiasis) can be a marker of elevated underlying cancer risk or shared risk factors. We also observe neighborhoods linking glaucoma and malignancy codes, in line with reports of slightly increased cancer risk among glaucoma patients [18].

#### (2) Inflammatory Bowel Disease (IBD) with spine disorders

Crohn’s disease and ulcerative colitis, collectively referred to as IBD are known to be associated with spine disorders, including ankylosing spondylitis and other inflammatory spondylopathies, that cause deformation of the spine. This relationship is also reflected in the embedding space.

#### (3) Arthritis with hearing loss and cataract

A shared neighborhood involving osteoarthritis, hearing loss, and cataract is consistent with age-related multimorbidity. A similar neighborhood involving rheumatoid arthritis may additionally reflect inflammation-related auditory impairment and treatment-mediated cataract risk (e.g., corticosteroid use). The embedding neighborhood may therefore combine age-driven and inflammation/treatment-driven pathways that generate similar co-occurrence signals.

##### Caveat

These qualitative examples are intended as interpretability signals rather than causal claims. First, t-SNE primarily preserves *local* structure and can distort global geometry; hence, we focus on nearest-neighbor patterns rather than large-scale distances. Second, frequency and healthcare-utilization effects can induce apparent comorbid neighborhoods (e.g., age-related conditions co-recorded during routine care). Systematic validation (e.g., quantitative neighborhood enrichment analyses, stratification by age/utilization, and clinician review) is left for future work.

### G Glossary of abbreviations

ALiBI: Attention with Linear Biases.
ATC: Anatomical Therapeutic Chemical (classification system for medications).
AUC: Area Under the Receiver Operating Characteristic Curve.
BERT: Bidirectional Encoder Representations from Transformers.
ICD: International Classification of Diseases.
CE: Cross-Entropy (loss).
CRC: Colorectal Cancer.
EHR: Electronic Health Record.
FFN: Feed-Forward Network.
GELU: Gaussian Error Linear Unit.
GP: General Practitioner (primary care).
hPCS: Hungarian Procedure Coding System.
ICD-10: International Classification of Diseases, 10th Revision.
ID: Identifier (e.g., token ID).
L1: Absolute error loss (mean absolute error).
L2: L2 regularization (squared ℓ _2_ penalty).
LN: Layer Normalization.
LR: Logistic Regression.
MLM: Masked Language Modeling.
MLP: Multi-Layer Perceptron.
PAD: Padding token (reserved ID for sequence padding).
MASK: Mask token (reserved ID for masking objectives).
PCA: Principal Component Analysis.
ROC: Receiver Operating Characteristic.
SHA-1: Secure Hash Algorithm 1 (hash function used for deterministic hashing).
Time2Vec: Continuous time embedding method based on periodic functions.
t-SNE: t-distributed Stochastic Neighbor Embedding.
UMAP: Uniform Manifold Approximation and Projection.

## Footnotes

1 We work on the administrative health records for billing purposes managed by the National Health Insurance Fund of Hungary (NHIF, Hungarian acronym: NEAK).

2 Code will be available at https://github.com/renyi-ai/HealthFormer.

3 Throughout the paper, ICD-10 codes are denoted as ICD.

4 In Hungary, a single public health insurer operates, the National Health Insurance Fund Hungary (Hungarian acronym: NEAK), to which all stakeholders of the healthcare delivery system submit the data required for reim-bursement on a monthly basis. In the context of the COVID–19 pandemic in 2020, a subset of patient flow data was transferred in anonymized form to the Health Services Management Training Centre of Semmelweis University responsible for the Health Data Utilization Team of the national pandemic management structures. There the interpretation, transformation, and linkage of these data enabled the establishment of a structured data warehouse suitable for research purposes.

5 We also built an interactive demo application to explore the learned ICD embedding space: it supports hierarchy-level views as well as a ICD search interface for interactive inspection of codes and their neighborhoods (https://ai.renyi.hu/healthcare/icd_embedding_visualizer/).

## Notes

### Competing Interest Statement

The authors have declared no competing interest.

### Author Declarations

Ethics Committee of the Hungarian Ministry of Interior gave ethical approval for this work (License No.: BM/14830-3/2024).

### Summary of Updates

- AI tool usage statement changed - Equal contribution statement added - Author order updated

